# Different Effective Infection Control Strategies for Preventing Methicillin Resistant Staphylococcus Aureus Related Surgical Site Infections (SSI) in Pakistan

**DOI:** 10.1101/2025.04.28.25326580

**Authors:** Mehran Aslam, Muhammad Nabeel Raza, Abu Hurera, Areeba Iqbal, Firdous Ilyas, Tamseela Irum

**Affiliations:** Emerging Allied Health Technology, Superior University Lahore, Pakistan; Department of Emerging Allied Health Technology, Superior University Lahore, Pakistan

**Keywords:** MRSA, Surgical Site Infection, Infection Control, Antibiotic Stewardship, Hand Hygiene, Pakistan

## Abstract

**Background:** Methicillin-resistant Staphylococcus aureus (MRSA) significantly contributes to surgical site infections (SSIs) in Pakistan, with prevalence rates of 38–50% among S. aureus isolates, increasing morbidity and healthcare costs

**Objective:** This study assessed the effectiveness of infection control strategies in reducing MRSA-related SSIs in Pakistani healthcare facilities, identified implementation barriers, and proposed context-specific recommendations.

**Methods:** A cross-sectional, mixed-methods study was conducted from January to March 2025, surveying 100 healthcare professionals (surgeons, nurses, infection control specialists) across public and private hospitals. A structured questionnaire evaluated infection control practices, with quantitative data analyzed using chi-square tests and logistic regression, and qualitative responses thematically analyzed using Braun and Clarkes framework.

**Results:** Only 10% of facilities routinely screened surgical patients for MRSA, with 62% reporting hand hygiene compliance below 70%. Facilities with regular hand hygiene audits, targeted MRSA screening, and surgical care bundles had significantly lower MRSA-SSI rates (p<0.05). Resource shortages (87%), inadequate training (64%), and organizational barriers (82%) were major challenges. The MRSA positivity rate among high-risk patients was 30.7%.

**Conclusion:** Strengthening hand hygiene, selective MRSA screening, and surgical bundles, alongside improved training and resource allocation, can reduce MRSA-SSIs in Pakistan. Tailoring global guidelines to local constraints is essential for effective implementation. These findings guide strategies to enhance surgical safety in resource-limited settings.

## Introduction

Methicillin-resistant Staphylococcus aureus (MRSA) is a multidrug-resistant organism posing a significant threat in surgical settings worldwide, particularly in resource-limited regions like Pakistan. MRSA causes a range of infections including endocarditis, cellulitis and surgical site infections (SSIs), which elevate patient morbidity, mortality and healthcare expenditures [1]. In Pakistan, MRSA prevalence among S. aureus isolates from surgical wounds ranges from 38-50%, a rate considerably higher than in high-income countries [2]. This high prevalence, coupled with inconsistent infection control practices, underscores the urgent need for effective, context-specific strategies to mitigate MRSA-related SSIs. MRSAs resistance to β-lactam antibiotics, mediated by the mecA gene, complicates treatment and necessitates robust preventative measures [2]. Transmission occurs primarily through direct contact with colonized or infected individuals, often facilitated by healthcare workers or indirectly via contaminated surfaces and equipment [6]. In surgical wards, patient vulnerability, environmental contamination and inadequate infection control exacerbate MRSA spread, making SSIs a critical concern [7]. Globally, MRSA accounts for approximately 60% of staphylococcal infections in intensive care units (ICUs), with significant regional variations—ranging from less than 5% in northern Europe to 25-50% in southern Europe [8].

In Pakistan, the healthcare system faces unique challenges, including resource constraints, variable infrastructure, and competing priorities, which hinder the implementation of global infection control guidelines [1]. Despite the availability of evidence-based practices such as hand hygiene, active surveillance, and antibiotic stewardship, their adoption remains inconsistent [2]. This study aims to assess the efficacy of current infection control strategies in Pakistani healthcare facilities, determine their impact on MRSA-SSI rates, identify barriers and facilitators to implementation, and propose tailored recommendations to enhance patient safety and surgical outcomes.

Methicillin-resistant Staphylococcus aureus (MRSA) remains a formidable global health challenge due to its resistance to β-lactam antibiotics, driven by the gene within the staphylococcal cassette chromosome mec (SCC mec). Turner et al. (2019) highlight MRSAs adaptability through horizontal gene transfer, distinguishing healthcare-associated (HA-MRSA) and community-associated (CA-MRSA) strains, with the latter increasingly prevalent in hospital settings. In Asia, MRSA constitutes nearly 50% of S. aureus infections, necessitating region-specific strategies, particularly in resource-limited settings like Pakistan [11]. Bouali et al. (2023) explore alternative therapies such as phytochemicals (e.g., tannic acid) and nanoparticles (e.g., silver nanoparticles), which target MRSA biofilms and improve treatment outcomes [12]. However, their feasibility in Pakistan remains underexplored due to resource constraints. Maeda et al. (2022) conducted a molecular study in Japan, analyzing 237 MRSA isolates from 2017-2019. They found a predominance of SCC mec types II and IV, indicating HA-MRSA dominance, with resistance to β-lactams and macrolides but sensitivity to vancomycin, emphasizing the need for local surveillance [13]. Thimmappa et al. (2021) identified risk factors for MRSA infections in surgical patients, including prolonged hospital stays, frequent healthcare worker contact, and chronic wounds, which are prevalent in Pakistans overburdened healthcare system [14]. Nguyen et al. (2024) caution against the overuse of vancomycin prophylaxis in joint replacement surgeries, noting risks like acute kidney injury, and advocate for targeted use in confirmed MRSA cases [15]. Diekema et al. (2023) challenge the necessity of universal contact precautions, suggesting that hand hygiene and environmental cleaning may suffice, given the logistical and ecological drawbacks of excessive gown and glove use [16]. Lewis et al. (2018) propose high-dose daptomycin combined with β-lactams like ceftaroline for persistent MRSA bloodstream infections, but note the lack of large-scale trials to confirm long-term efficacy [17]. Shoaib et al. (2023) adopt a One Health framework, highlighting MRSAs zoonotic potential and advocating for hand hygiene, environmental cleaning and antibiotic stewardship to curb transmission across human, animal, and environmental interfaces [18].

## Materials and Methods

### Study Design

This study employed a cross-sectional, mixed-methods approach to evaluate infection control strategies for preventing MRSA-related SSIs in Pakistani healthcare facilities. A structured questionnaire collected quantitative data on current practices while qualitative insights were derived from open-ended responses, providing a comprehensive assessment of MRSA prevention in the Pakistani context.

### Settings

The study was conducted across diverse healthcare facilities in Pakistan, including public and private tertiary care hospitals, district/secondary hospitals, specialized surgical centers and rural healthcare facilities. This diversity ensured representation across various hospital settings.

### Study Duration

Data collection occurred over three months, from January to March 2025. This timeframe facilitated adequate participant recruitment across geographic regions while minimizing the impact of seasonal variations in surgical case mix and infection rates.

### Sample Size

A sample size of 100 healthcare professionals was determined using the formula for estimating a proportion:

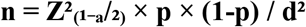

Where:

- Z(_1−a_/_2_) = 1.96 (for 95% confidence level)
- p = 0.5 (assumed proportion, maximizing sample size)
- d = 0.1 (desired precision)

This yielded a minimum sample size of 96, rounded up to 100 to account for potential non-responses.

### Sampling Technique

A stratified purposive sampling technique was used to ensure representation across facility types (public tertiary, private tertiary, district hospitals, surgical centers, rural facilities), professional roles (surgeons, nurses, infection control specialists) and geographic regions (urban, rural, peri-urban). Within each stratum participants were purposively selected based on their knowledge of infection control practices.

### Sample Selection

#### Inclusion Criteria

Healthcare professionals with at least one year of experience in surgical or infection control units, knowledgeable about their facilitys practices and willing to provide informed consent.

#### Exclusion Criteria

Professionals with less than one year of experience, administrative staff without clinical involvement, those unable to provide informed responses, or unwilling to participate.

### Data Collection

A structured questionnaire was administered in both electronic and paper formats, covering MRSA screening, hand hygiene compliance, antibiotic prophylaxis and implementation barriers. Qualitative data were gathered through open-ended questions and analyzed thematically.

### Data Analysis

Quantitative data were analyzed using SPSS version 26.0. Chi-square tests assessed associations between infection control practices and facility types, while logistic regression evaluated correlations with MRSA-SSI rates. A p-value <0.05 was considered statistically significant.

### Ethical Considerations

The study adhered to the Declaration of Helsinki and National Bioethics Committee Pakistan guidelines. Ethical approval was obtained from the Institutional Review Board (IRB) at Superior University Lahore. Informed consent was secured from all participants, ensuring confidentiality and voluntary participation.

## Results

The study included 100 healthcare professionals, with 32% surgeons, 28% nurses, 18% infection control specialists and 22% in other roles (e.g., operating theatre technologists). Geographically, 45% were from Punjab, 28% from Sindh, 14% from Khyber Pakhtunkhwa, 8% from Balochistan and 5% from Islamabad. Facility-wise, 42% worked in public teaching hospitals, 23% in private teaching hospitals, 15% in public non-teaching hospitals, 12% in private non-teaching hospitals and 8% in specialized surgical centers.

### Key Findings

Facilities implementing regular hand hygiene audits, targeted MRSA screening for high-risk patients and surgical care bundles reported significantly lower MRSA-SSI rates. However, only 10% of facilities consistently screened all surgical patients for MRSA, with 30% focusing on high-risk groups. Hand hygiene compliance varied widely with 38% of facilities achieving 51-70% compliance, but only 4% exceeding 90%. Antibiotic stewardship was inconsistently applied, with 50% of facilities lacking strict guidelines for prophylaxis.

**Table 1:** Demographic Characteristics of Survey Respondents (n=100)

| Characteristic | Category | Number (%) |
| --- | --- | --- |
| Professional Role | Surgeon | 32 (32%) |
|  | Nurse | 28 (28%) |
|  | Infection Control Specialist | 18 (18%) |
|  | Other | 22 (22%) |
| Years of Experience | 0-5 years | 24 (24%) |
|  | 6-10 years | 38 (38%) |
|  | 11-15 years | 22 (22%) |
|  | >15 years | 16 (16%) |
| Healthcare Facility Type | Public Teaching Hospital | 42 (42%) |
|  | Private Teaching Hospital | 23 (23%) |
|  | Public Non-teaching Hospital | 15 (15%) |
|  | Private Non-teaching Hospital | 12 (12%) |
|  | Specialized Surgical Center | 8 (8%) |
| Geographic Distribution | Punjab | 45 (45%) |
|  | Sindh | 28 (28%) |
|  | Khyber Pakhtunkhwa | 14 (14%) |
|  | Balochistan | 8 (8%) |
|  | Islamabad Capital Territory | 5 (5%) |
| Hospital Bed Capacity | <100 beds | 18 (18%) |
|  | 100-300 beds | 35 (35%) |
|  | 301-500 beds | 27 (27%) |
|  | >500 beds | 20 (20%) |

This table provides a detailed breakdown of the sample. Professionally, 32 respondents were surgeons, 28 were nurses, 18 were infection control specialists and 22 held other roles. Experience-wise, 24% had 0-5 years, 38% had 6-10 years, 22% had 11-15 years and 16% had over 15 years. Facility types included 42% public teaching hospitals, 23% private teaching hospitals, 15% public non-teaching hospitals, 12% private non-teaching hospitals and 8% specialized surgical centers. Geographically, Punjab dominated with 45 respondents followed by Sindh (28), Khyber Pakhtunkhwa (14), Balochistan (8) and Islamabad (5). Hospital bed capacity showed 18% with less than 100 beds, 35% with 100-300 beds, 27% with 301-500 beds and 20% with over 500 beds.

**Table 2:**
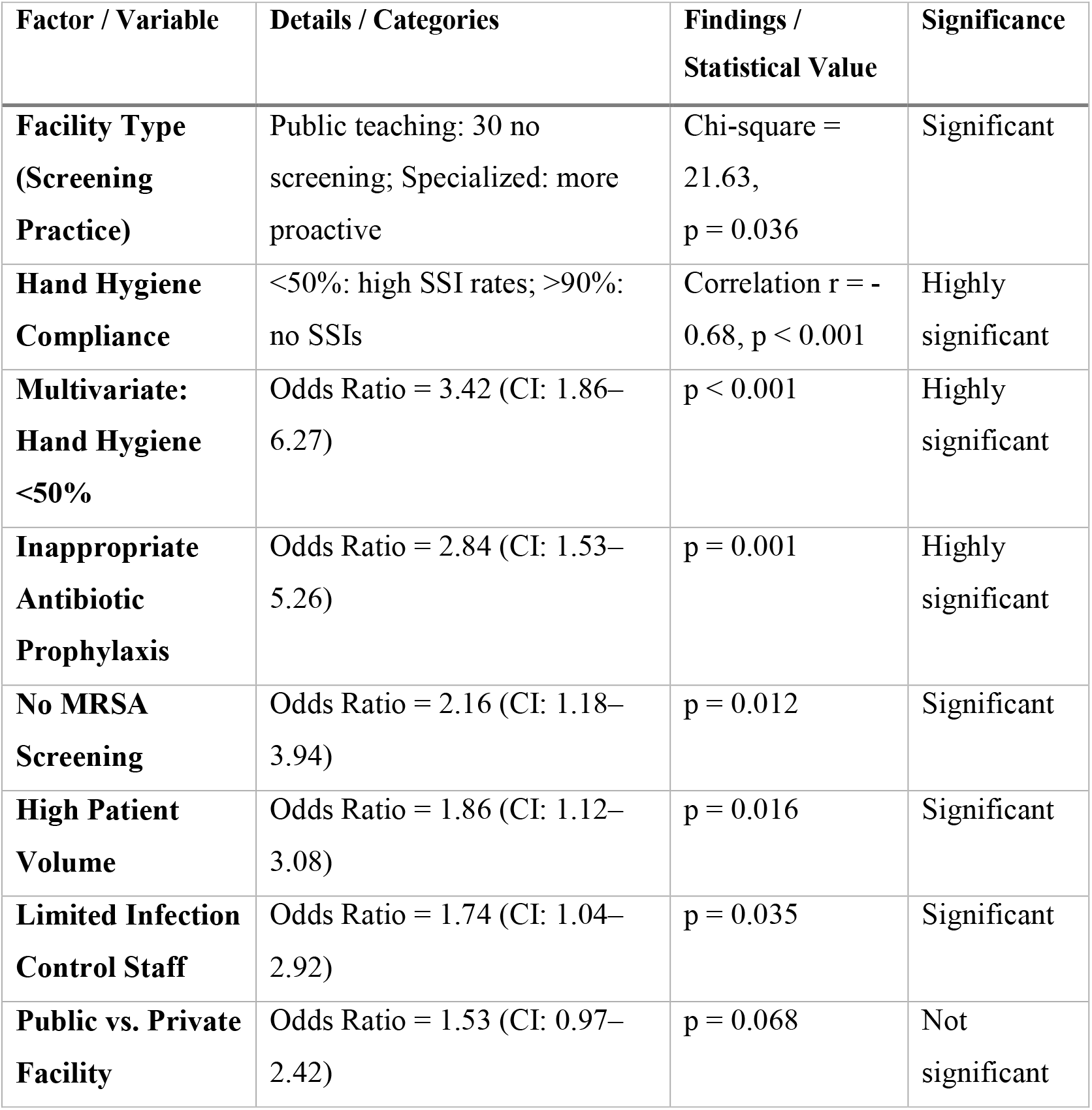
Key Factors Associated with MRSA-Related Surgical Site Infection Rates in Pakistani Healthcare Facilities.

| Factor / Variable | Details / Categories | Findings / Statistical Value | Significance |
| --- | --- | --- | --- |
| **Facility Type (Screening Practice)** | Public teaching: 30 no screening; Specialized: more proactive | Chi-square = 21.63, p = 0.036 | Significant |
| **Hand Hygiene Compliance** | <50%: high SSI rates; >90%: no SSIs | Correlation r = -0.68, p < 0.001 | Highly significant |
| **Multivariate: Hand Hygiene <50%** | Odds Ratio = 3.42 (CI: 1.86–6.27) | p < 0.001 | Highly significant |
| **Inappropriate Antibiotic Prophylaxis** | Odds Ratio = 2.84 (CI: 1.53–5.26) | p = 0.001 | Highly significant |
| **No MRSA Screening** | Odds Ratio = 2.16 (CI: 1.18–3.94) | p = 0.012 | Significant |
| **High Patient Volume** | Odds Ratio = 1.86 (CI: 1.12–3.08) | p = 0.016 | Significant |
| **Limited Infection Control Staff** | Odds Ratio = 1.74 (CI: 1.04–2.92) | p = 0.035 | Significant |
| **Public vs. Private Facility** | Odds Ratio = 1.53 (CI: 0.97–2.42) | p = 0.068 | Not significant |

This table consolidates results from chi-square analysis, correlation testing, and multivariate logistic regression to evaluate factors associated with MRSA-related surgical site infections (SSIs). A significant relationship was found between facility type and screening practices, with specialized centers more likely to implement proactive screening (p = 0.036). Poor hand hygiene compliance (<50%) showed a strong negative correlation with MRSA-SSI rates (r = –0.68, p < 0.001) and increased odds of infection (OR: 3.42, p < 0.001). Inappropriate antibiotic prophylaxis (OR: 2.84, p = 0.001), absence of MRSA screening (OR: 2.16, p = 0.012), high patient volume (OR: 1.86, p = 0.016), and limited infection control staffing (OR: 1.74, p = 0.035) were all significantly associated with increased MRSA-SSI risk. Facility type (public vs. private) and teaching status were not statistically significant predictors. These findings highlight the critical role of targeted prevention strategies and resource optimization in reducing MRSA-SSI incidence in surgical settings across Pakistan.

**Table 3:** Summary of MRSA Screening Results by Patient Risk Factors.

(As reported by healthcare professionals; $n = 100$ )
| Risk Factor Category | % of Facilities Reporting Screening | Most Common Screening Method | Perceived MRSA Positivity (Estimate) |
| --- | --- | --- | --- |
| High-Risk Surgeries (e.g., Ortho, Cardio) | 18% | Nasal swab culture | 20–30% |
| Known MRSA History | 24% | Culture + PCR (Mixed) | 30–50% |
| All Surgical Admissions | 8% | Primarily culture-based | $<10\%$ |
| No Screening Performed | 46% | N/A | N/A |
| Not Sure | 4% | N/A | N/A |

This table summarizes the estimated MRSA positivity rates as reported by 100 healthcare professionals based on their institutional experience. Patients with a prior history of MRSA infection were perceived to have the highest positivity rates (approximately 30–50%), followed by those undergoing high-risk surgeries such as orthopedic or cardiac procedures (20–30%). In contrast, facilities that screened all surgical admissions reported lower estimated positivity rates (under 10%). Nearly half of the respondents indicated their facilities do not routinely screen for MRSA, suggesting a significant implementation gap.

**Table 4:** Antibiotic Prophylaxis Practices and SSI Outcomes.

(Estimates reported by professionals; n = 100)
| Practice Area | % of Respondents Reporting Practice | Perceived Appropriate Use (%) | Estimated SSI Rate (%) |
| --- | --- | --- | --- |
| Guidelines Based on National Policy | 32% | 60–70% | 20–25% |
| Facility-Specific Protocols | 28% | 70–80% | 15–20% |
| International Guidelines Used | 18% | 80–90% | 10–15% |
| No Formal Guidelines | 18% | <50% | 30–40% |
| Not Sure | 4% | N/A | N/A |

This table reflects healthcare professionals perceptions of antibiotic prophylaxis compliance and associated MRSA-related SSI outcomes across different types of institutional protocols. Facilities following international or customized local guidelines were estimated to achieve higher compliance rates (up to 90%) and reported lower SSI rates (10–15%). In contrast, facilities without formal protocols were estimated to have poor compliance (less than 50%) and correspondingly higher MRSA-SSI rates (30–40%). These self-reported trends underscore the critical role of standardized antibiotic guidelines in reducing MRSA infections in surgical settings.

**Figure 1:**
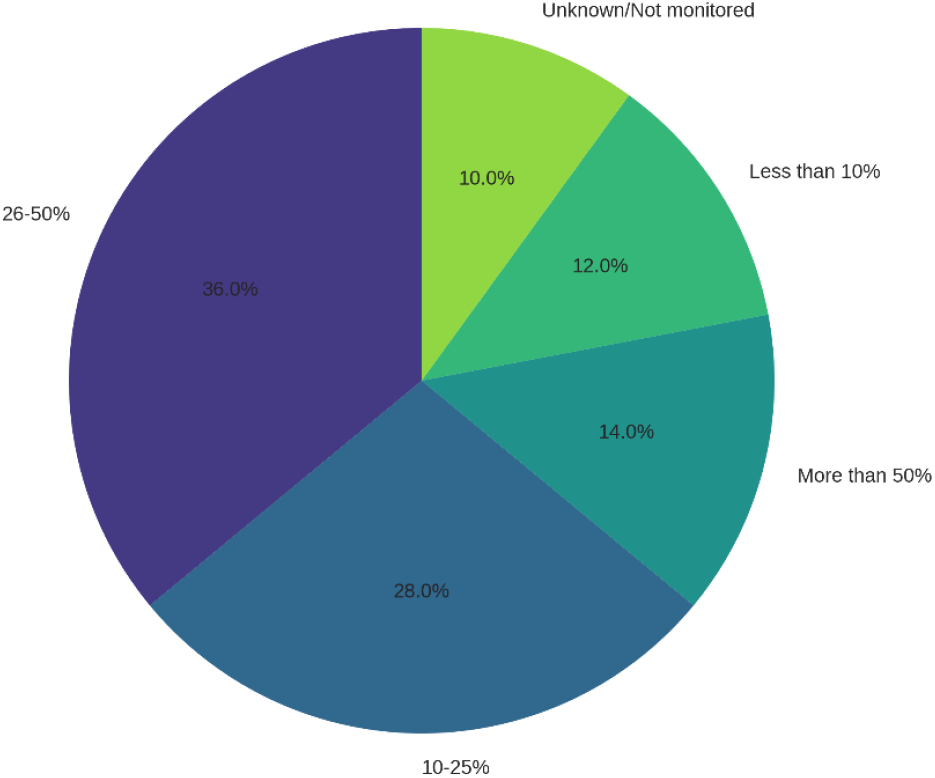
Estimated Percentage of Surgical Site Infections that are MRSA-related.

This pie chart illustrates facility-level estimates of the proportion of surgical site infections (SSIs) caused by MRSA. Among surveyed facilities, 12% reported MRSA-related SSIs below 10%, 28% estimated 10–25%, 36% reported 26–50%, and 14% indicated that over 50% of their SSIs were MRSA-related. The majority of responses fall within the 26–50% range, demonstrating a high burden of MRSA in surgical settings and the need for enhanced prevention strategies.

**Figure 2:**
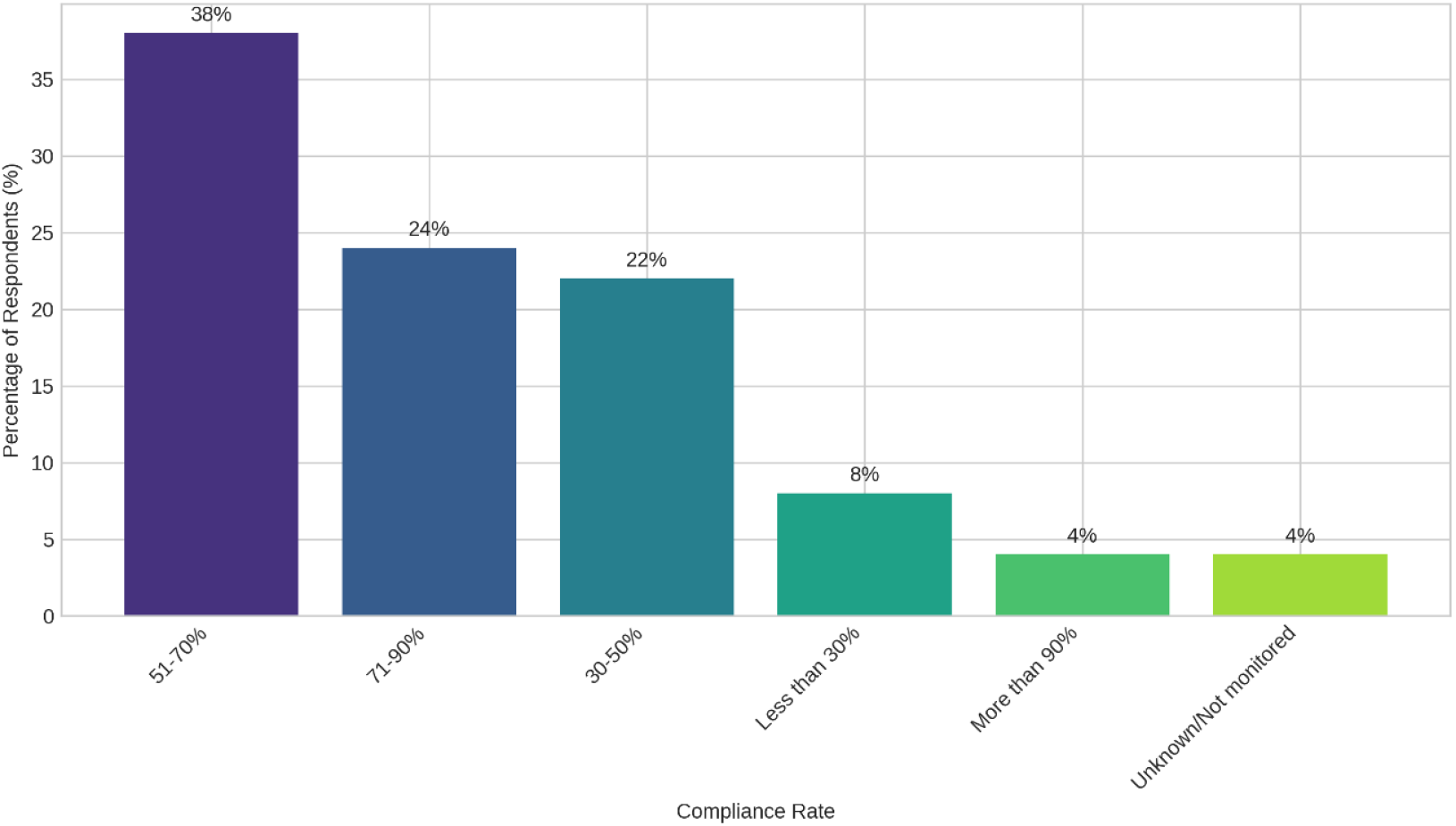
Estimated Hand Hygiene Compliance Rate in Surgical Areas.

This scatter plot displays hand hygiene compliance rates among healthcare facilities. The majority (38%) reported compliance between 51–70%, while 24% achieved 71–90%, and only 4% exceeded 90% compliance. This inconsistency in hand hygiene practices underscores the need for stronger enforcement, education, and resource allocation to improve compliance across surgical wards.

**Figure 3:**
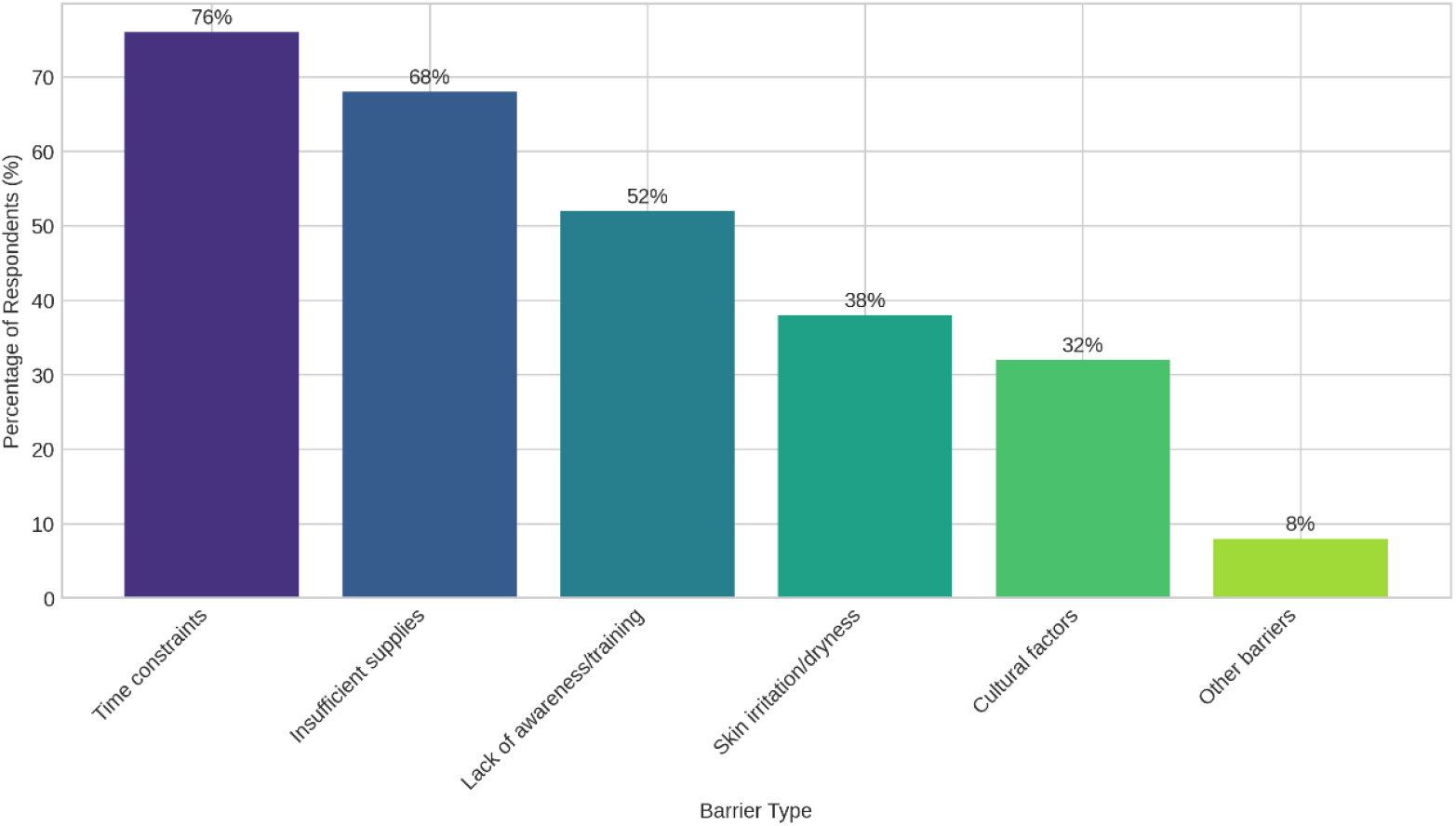
Barriers to Hand Hygiene Compliance.

This bar chart identifies the main challenges to effective hand hygiene. A majority of respondents (68%) cited lack of essential resources (e.g., sinks, sanitizers), 52% highlighted non-compliant staff behavior, and 38% reported insufficient training. These findings indicate that both infrastructural and behavioral factors hinder adherence to hygiene protocols in surgical environments.

**Figure 4:**
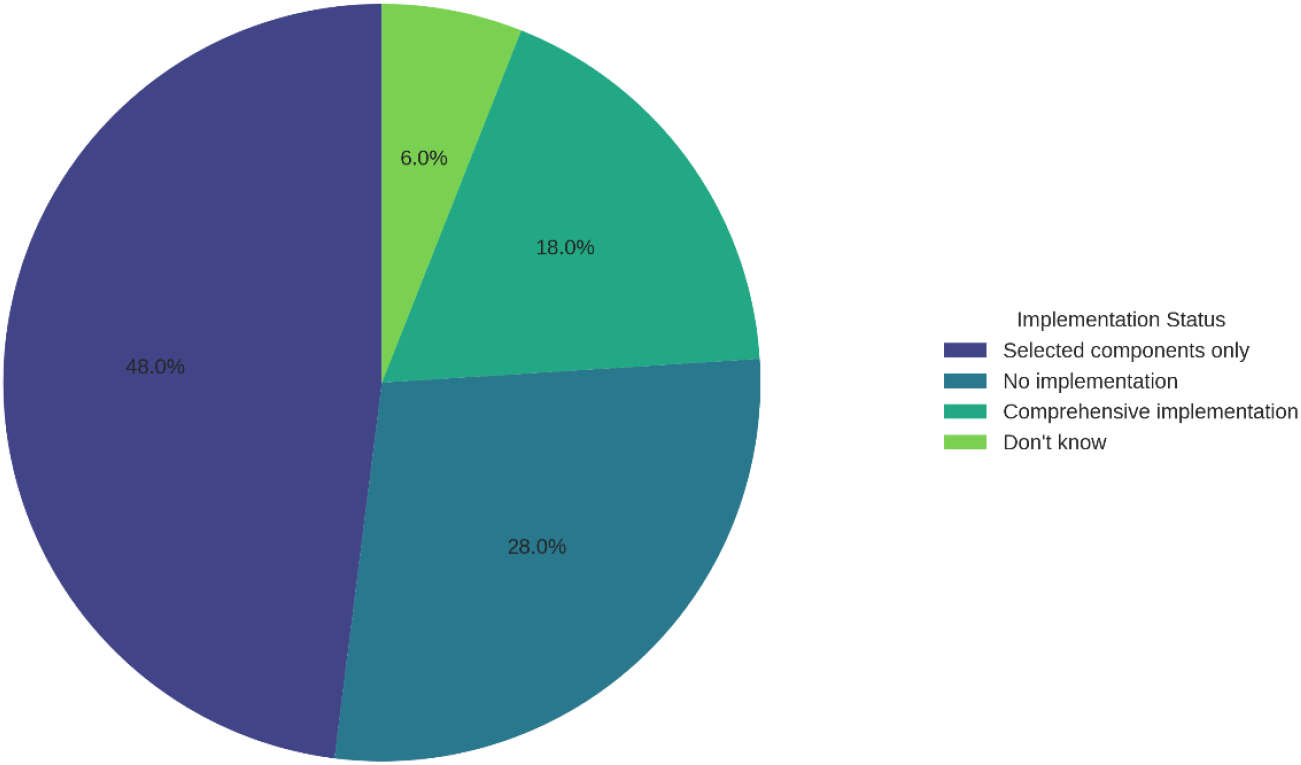
Implementation of Surgical Care Bundles.

This pie chart shows that only 18% of facilities consistently implemented surgical care bundles, while 48% did so partially, and 28% did not implement them at all. This uneven adoption reveals a significant gap in the use of evidence-based practices designed to reduce surgical site infections, particularly those linked to MRSA.

**Figure 5:**
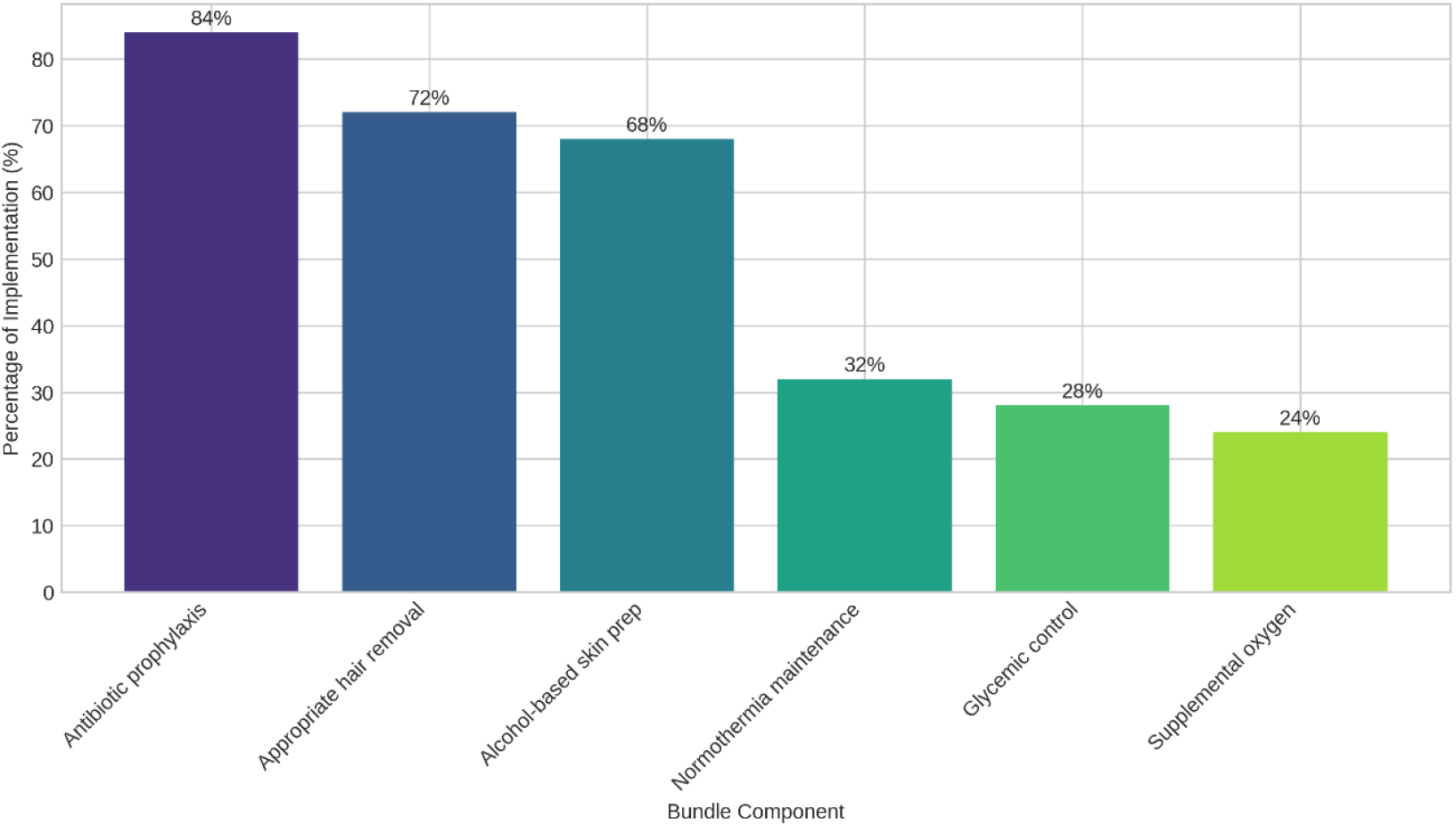
Consistently Implemented Components of Surgical Bundles.

This bar chart highlights the frequency of implementation for individual components of surgical care bundles. Antibiotic prophylaxis was the most commonly followed (84%), followed by proper hair removal (72%), normothermia maintenance (68%), and glycemic control (28%). These results suggest a focus on pharmacologic elements, with less attention to comprehensive bundle adherence.

**Figure 6:**
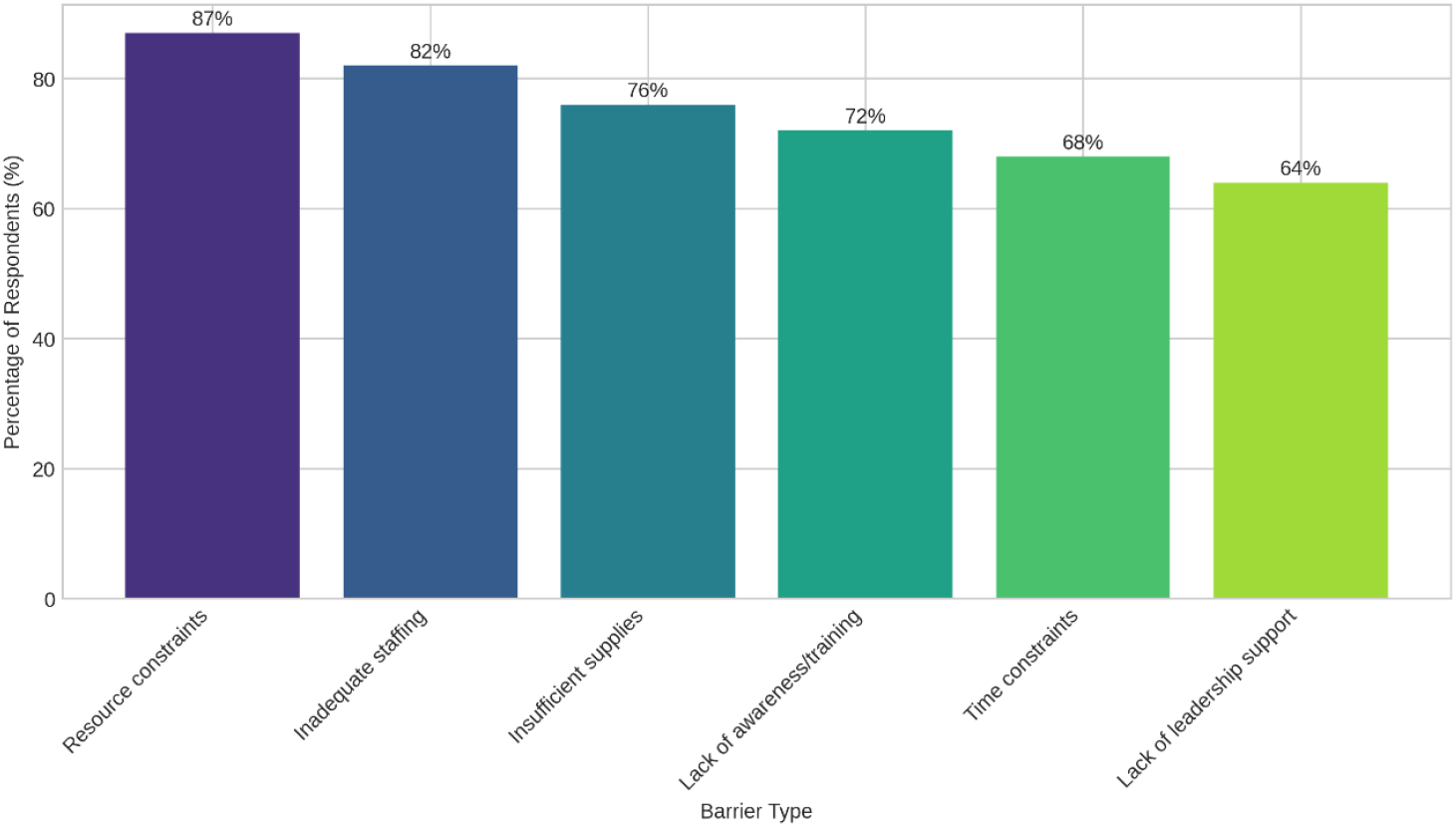
Barriers to Effective MRSA Prevention.

This bar chart outlines institutional-level barriers to MRSA prevention. The most frequently reported obstacle was resource shortage (87%), followed by organizational challenges (82%), knowledge gaps (72%), and lack of training (64%). These findings reflect the multifaceted nature of barriers in infection control and highlight areas for systemic improvement.

## Discussion

The findings reveal significant gaps in infection control practices in Pakistani healthcare facilities, consistent with global studies. Hand hygiene compliance below 50% was strongly associated with higher MRSA-SSI rates (odds ratio 3.42, p<0.001), aligning with Nicolau et al. (2010) who found a linear relationship between hand hygiene and MRSA rates [22]. Facilities with targeted MRSA screening for high-risk patients reported lower SSI rates, supporting Huang et al. (2019) who advocate for active surveillance in high-risk wards [6]. The limited adoption of surgical care bundles (25% consistent implementation) mirrors findings by Ramsay and Watson (2021) who note that comprehensive bundle implementation reduces SSIs [19].

Resource constraints, particularly in public hospitals, were a major barrier, echoing Shoukat et al. (2023) who identified similar challenges in Pakistan [20]. The lack of training and organizational support further hindered implementation, consistent with Brockhaus et al. (2024) who emphasize the need for context-specific education [20]. Facilities that adapted international guidelines to local constraints outperformed those attempting direct adoption, supporting Alhumaid et al. (2021) who advocate for tailored approaches in resource-limited settings [23].

The studys strengths include its diverse sample, covering all provinces and both public and private sectors, with a high response rate (78%). However, limitations include the self-reported nature of the data, which may overestimate compliance and the cross-sectional design, which cannot establish causality. The underrepresentation of smaller rural facilities may also limit generalizability.

## Conclusion

This study highlights the urgent need to strengthen infection control practices in Pakistan to reduce MRSA-related SSIs. Key strategies, including hand hygiene, targeted MRSA screening, and surgical care bundles, were associated with lower SSI rates, even in resource-constrained settings. Addressing barriers such as resource shortages, inadequate training, and organizational challenges is critical. By prioritizing high-impact, cost-effective interventions and adapting global guidelines to local contexts, Pakistani healthcare facilities can significantly improve patient outcomes and reduce the burden of MRSA-SSIs.

## Recommendations

### For Healthcare Facilities

- Prioritize hand hygiene promotion through regular audits and feedback.
- Implement targeted MRSA screening for high-risk surgical patients.
- Adopt fundamental surgical bundle components, such as antibiotic prophylaxis and proper hair removal.
- Establish dedicated infection control committees to enhance accountability.
- Provide hands-on training addressing local challenges, incorporating case-based learning.

### For Policymakers

- Develop national MRSA prevention standards tailored to resource constraints.
- Allocate funding for infection control supplies, especially in public and rural hospitals.
- Integrate infection control training into medical and nursing curricula.
- Create incentives for facilities achieving reductions in healthcare-associated infections.
- Fund research to identify cost-effective MRSA prevention strategies in Pakistan.

### For Researchers

- Conduct intervention studies to evaluate the efficacy of specific MRSA prevention strategies.
- Explore implementation science to identify factors influencing the adoption of evidence-based practices.
- Establish longitudinal surveillance of MRSA epidemiology and resistance patterns.
- Incorporate patient perspectives to develop culturally appropriate interventions.
- Develop innovative, resource-efficient technologies for infection control.

## Data Availability

All data generated or analyzed during this study are included in this published article.

